# Statistical analysis plan for the “Clinical effectiveness of peripheral intravenous device selection and insertion by a vascular access specialist” (The SELECT Study)

**DOI:** 10.64898/2025.12.03.25341353

**Authors:** Maharshi Patel, Nicole Marsh, Asmaa El-Heneidy, Catherine O’Brien, Emily Larsen, Amanda Corley, Robert S. Ware

## Abstract

The “Clinical effectiveness of peripheral intravenous device selection and insertion by a vascular access specialist” (The SELECT Study)” is a single-centre two-arm superiority randomised clinical trial (RCT). This RCT aims to compare peripherally placed IV device selection and insertion by a Vascular access specialist with a generalist inserter.

The purpose for this document is to minimise bias and ensure transparency and internal validity for the findings of the trial, by defining and making publicly available the analysis approach prior to reviewing or analysing trial data. The statistical analysis plan will inform analysis and reporting of the main effectiveness findings of the trial. It provides a detailed description of the primary and secondary trial outcomes and the methods for statistical comparison.

## 1.0 Administrative information

### 1.1 Purpose

The SELECT study is a two-armed randomised controlled trial (RCT) that seeks to determine the superiority of vascular access specialists (VAS) compared with any other clinician on peripherally placed IV device selection and insertion.

The purpose for this document is to minimise bias and ensure transparency and internal validity for the findings of the trial, by defining and making publicly available the analysis approach prior to reviewing or analysing trial data. This statistical analysis plan (SAP) will inform analysis and reporting of the main effectiveness findings of the trial. It provides a detailed description of the primary and secondary trial outcomes and the methods for statistical comparison.

### 1.2 Study identifiers

- Protocol document: v1.1, 23/03/2023.
- ANZCTR Registration Record: ACTRN12623000489695

## 2.0 Background

Peripheral intravenous catheters (PIVCs) are the most frequently used vascular access device in Australia, with Queensland Health spending $41 million purchasing over 2.6 million PIVCs in 2016 alone^1^. However, around half of all PIVCS fail before the completion of therapy^2,3^ leading to: delays in essential intravenous (IV) treatments (e.g., antibiotics, chemotherapy); repetitive and painful reinsertion procedures (average re-insertion delay, 50 hours^4^); and increased healthcare costs (staff time/consumables)^4^. Consequently, improved insertion practices and workforce models would lead to significant cost savings^5^.

Currently, peripheral IV device choice consistently defaults to short-PIVCs (<4cms) to meet most patients’ IV treatment needs^6^. This is largely due to the current workforce model in which most PIVCs inserters are junior doctors and nurses inserting PIVCs within their existing roles (‘generalist’ approach)^5^, however these staff often have limited knowledge and training of 1) IV device options (short/long PIVCs, midline catheters [MC]), 2) correct device selection for treatment needs, and 3) optimal insertion techniques, particularly in patients with difficult IV access (DIVA)^4,7^. Alternate devices to the short-PIVC such as long-PIVCs (4-6.3cms) and MCs (device tip sits at the axilla fold) are available and these longer devices allow greater catheter length to reside within the vein, decreasing risk of catheter dislodgement and failure^8,9^. Recent observational data has shown extended dwell times for longer devices (7.7–16.4 days)^10^, compared to short-PIVCs (2.4-3.5 days)^4,11^.

Over the last decade, PIVC insertion teams made up of Vascular Access Specialists (VAS) (predominately nurses, with advanced PIVC insertion and maintenance skills), have been disbanded in many hospitals due to budgetary constraints^12,13^. The ‘generalist’ approach has been adopted instead. However, this workforce has not been supported in literature, with recent studies (including one early-stage feasibility randomised controlled trial (RCT) supported by the RBWH Foundation^2^) finding lower first-time insertion success, higher rates of complications and poorer patient satisfaction with generalist inserters compared to a VAS^2,5,7,12^. The ‘generalist’ workforce model puts pressure on already-stretched frontline clinicians, particularly junior medical officers in the after-hours space. A recent RBWH audit of after-hours referrals, led by CI Peters, revealed significant healthcare inefficiencies^14^. In total, 561 PIVC insertion referrals (22% of which were patients with DIVA), were made to junior medical staff over a 3-month period. This resulted in: 48% of insertions for patients with DIVA being referred to another inserter, half of whom were senior medical staff; high likelihood of multiple failed insertion attempts (with less than half of DIVA patients having a PIVC successfully inserted after-hours); delays to IV treatment; and poor patient satisfaction^14^.

Inherent inefficiencies in the current generalist workforce model are not sustainable: more efficient, innovative service provision is urgently needed. We plan to conduct a rigorous RCT comparing clinical outcomes of a dedicated VAS workforce model with the current generalist inserter approach. A qualitative study will be conducted concurrently, to explore the current state of the generalist inserter approach (including opportunity cost and larger healthcare institution implications).

## 3.0 Study synopsis

This study is a single-centre two-arm superiority, controlled study with a 1:1 (VAS: generalist) randomisation of participants.

### 3.1 Study objectives

#### 3.1.1 Primary objective

The primary objective is to compare the clinical effectiveness of peripheral intravenous (IV) device selection and insertion by (i) a VAS compared with (ii) any clinician (generalist model; standard practice).

### 3.2 Eligibility criteria

#### 3.2.1 Inclusion criteria Participants are included in the study if they

- are ≥18 years of age
- are requiring peripherally compatible IV therapy for ≥ 24hours
- have informed written consent

#### 3.2.2 Exclusion criteria

Participants are excluded from the study if they:

- are receiving end-of-life care
- have limited English, without appropriate interpreter
- have previous study enrolment

### 3.3 Outcomes

#### 3.3.1 Primary outcome

The primary clinical outcome is Peripherally-placed IV device failure: a composite of infiltration/extravasation, blockage/occlusion (with/without leakage), phlebitis, (defined as: pain score equal to or greater than 2 alone; OR two or more signs/symptoms of either: pain/tenderness; erythema/redness; swelling or palpable cord), thrombosis, dislodgement (complete/partial) or infection (laboratory-confirmed local/bloodstream infection).

#### 3.3.2 Secondary outcomes

1. Infiltration/extravasation (defined as the movement of IV fluid/vesicant into surrounding tissue, with or without resulting tissue damage)
2. Blockage/occlusion (with/without leakage defined as: PIVC will not infuse/flush or leakage occurs when fluids infused/flushed.)
3. Phlebitis (as defined above)
4. Thrombosis (defined as thrombosed vessel related to the allocated IV device confirmed by ultrasound/venographic imaging)
5. Dislodgement (defined as Partial - change in device length at insertion site, resulting in device removal. Complete - device completely leaves the vein.
6. Local Infection (laboratory-confirmed local infection defined as: Localised peripheral IV site infection (without bloodstream infection) as per NHSN 2023 CVS-VASC criteria.
7. Bloodstream Infection (Laboratory Confirmed Bloodstream Infection as defined by NHSN 2023 criteria. For Central Line: in place for 2 days or more. PIVC and Midline Catheter: must also meet requirement of above laboratory-confirmed local infection.
8. First insertion success
9. Failed insertion (complete failure to insert any device)
10. Number of failed insertion attempts (needle punctures)
11. Peripherally placed IV device dwell-time (hours) from device insertion to removal
12. Number and type of devices needed to complete IV treatment (‘completion’: hospital discharge or no IV device required for 7 days)
13. Patient-reported pain at device insertion (0-10 rating scale)
14. Patient-reported satisfaction with IV device.

### 3.4 Interventions

Participants randomised to either:

1. Device selection and insertion by a VAS (short-PIVC, long-PIVC, MC) or PICC recommendation.
2. Device selection and insertion by a generalist inserter, including the option to escalate to another clinician or service, as per current hospital practice.

### 3.5 Setting

The RCT will be undertaken in the medical and surgical wards of the Royal Brisbane and Women’s Hospital.

### 3.6 Randomisation

Randomisation was via a central, web-based service (1:1), where participants received a device inserted by either a generalist clinician or VAS, with randomly varied block sizes of four and six, and with allocation concealment until commencement of study procedures.

### 3.7 Sample size

Using pilot data, we expected peripheral IV device failure to fall from 54% (generalist inserters)^2^ to less than 34% (VAS inserters)^15^, an absolute difference of 20%. This is equivalent to a decrease in odds of 54% (i.e., odds ratio < 0.44). To detect a statistically significant between-group difference (alpha=0.05, 80% power, 5% attrition), 196 participants (93 per arm + 10 for attrition) were needed. This ensured ≥30 episodes of failure for robust estimates of costs in the economic analysis.

## 4.0 Statistical analysis

### 4.1 General Principals

#### 4.1.1 Intention-to-treat and per-protocol analyses

All participants who are randomised and have evaluable data for the endpoints under investigation, will be analysed in the study arm to which they were randomised. Participants who are recruited, and then whose insertion is cancelled, or who withdraw consent will not be included in the intention-to-treat sample.

For the primary outcome, an instrumental variables per-protocol analysis will be performed to identify the causal effect of the intervention in those who receive it. For this per-protocol analysis, randomisation will be the independent variable and treatment received will be the exposure variable. If there is no treatment non-compliance this analysis will not be conducted

The number of missing observations for each primary and secondary outcome will be reported by study group. If the proportion of missing observations is considerable, consideration will be given to the possible introduction of bias for the between-group comparisons. The cause of any missing data will be assessed, and sensitivity analysis to investigate the potential impact of missing data will be undertaken using multiple imputation techniques if appropriate.

#### 4.1.2 Presentation of results

Continuous data will be summarized descriptively using either mean and standard deviation (SD) or median and interquartile range (IQR), depending on the distribution of the variable of interest. Categorical data will be presented as frequencies and percentages.

#### 4.1.3 Level of significance

A significance level of alpha = 0.05 (two-tailed) will be used to evaluate statistical significance for the primary outcome. Secondary outcomes will be treated as exploratory.

#### 4.1.4 Statistical software

Trial data will be exported from the REDCap database to Stata statistical software v16 (StataCorp, College Station, TX, USA) for analysis.

### 4.2 Description of demographic and baseline characteristics

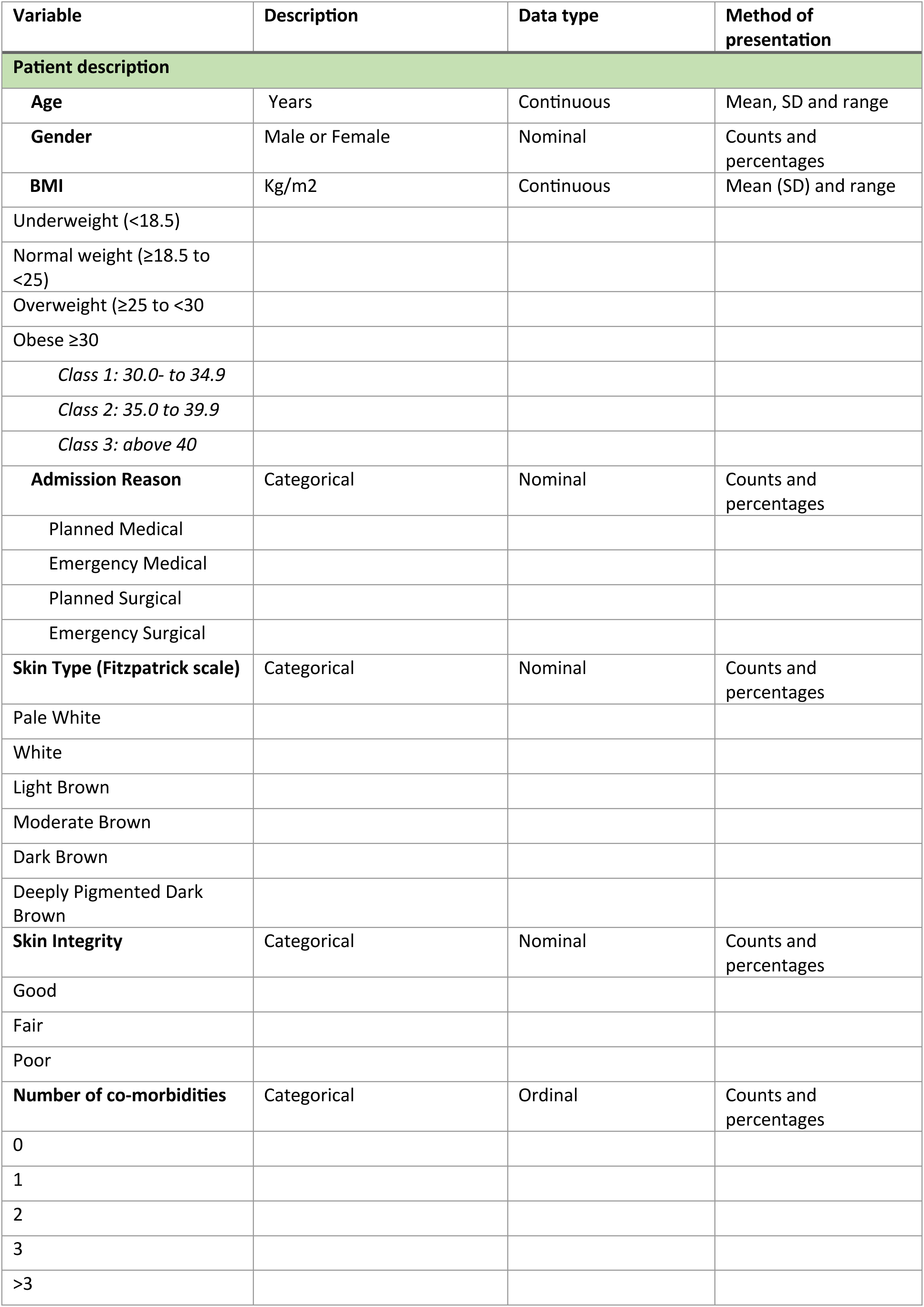
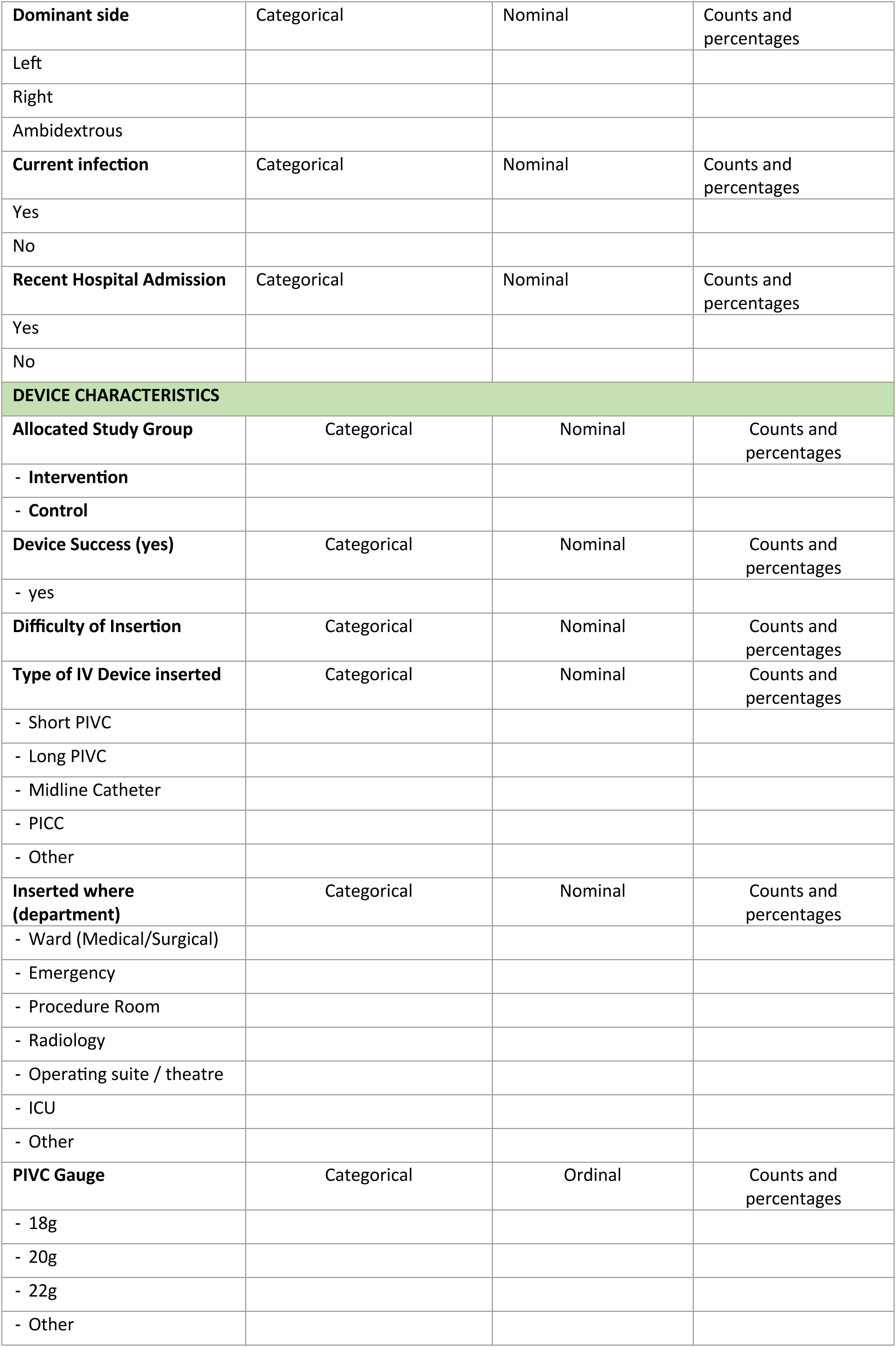
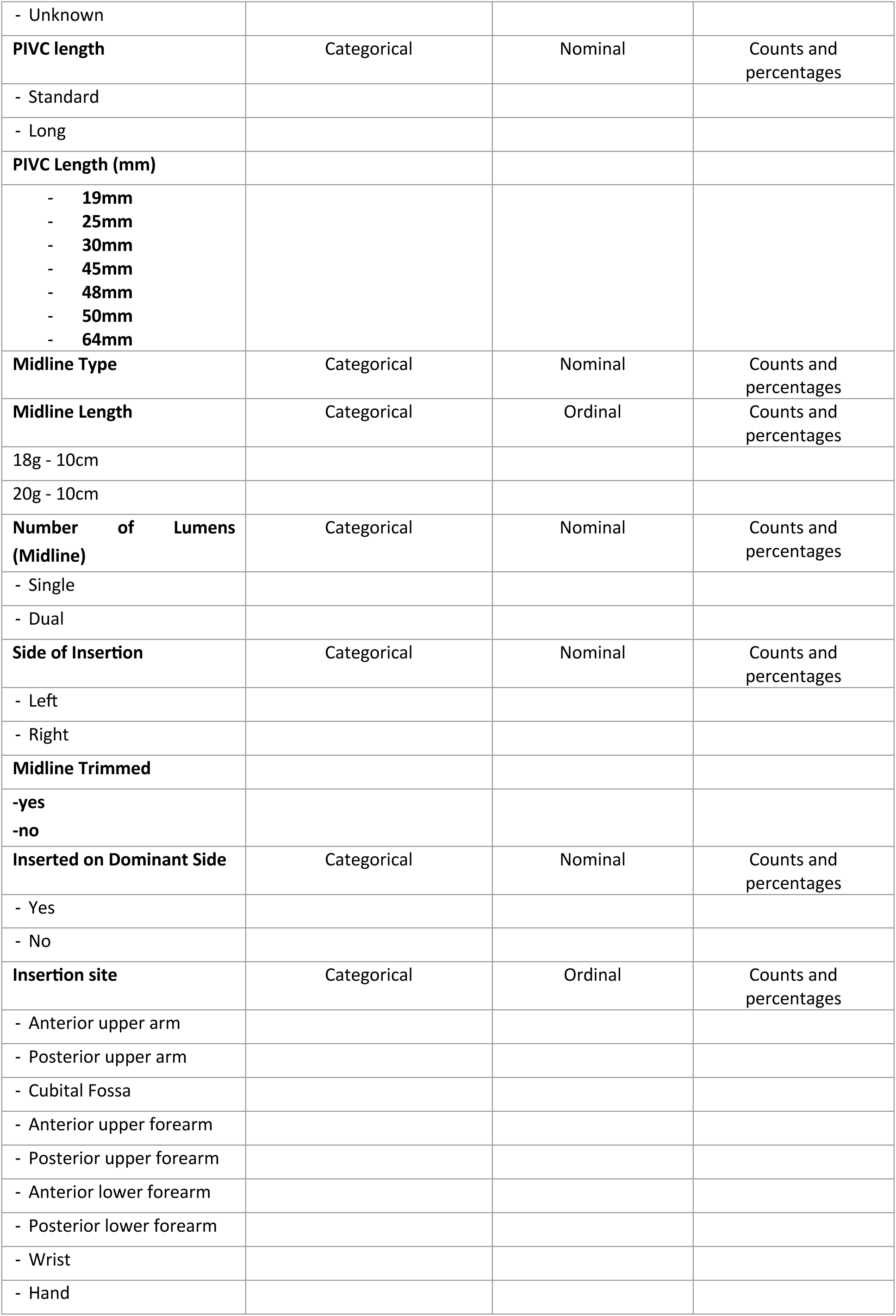
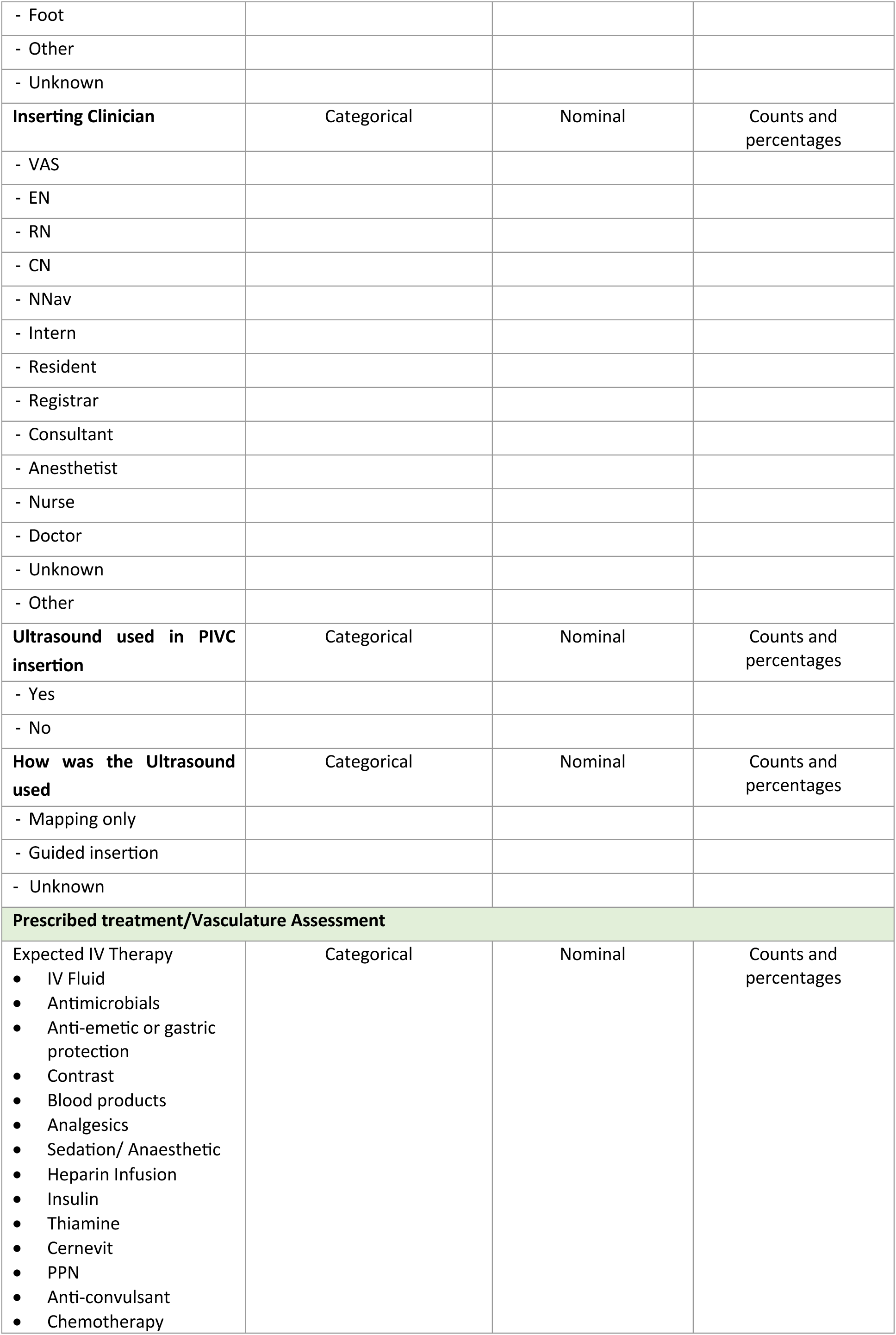
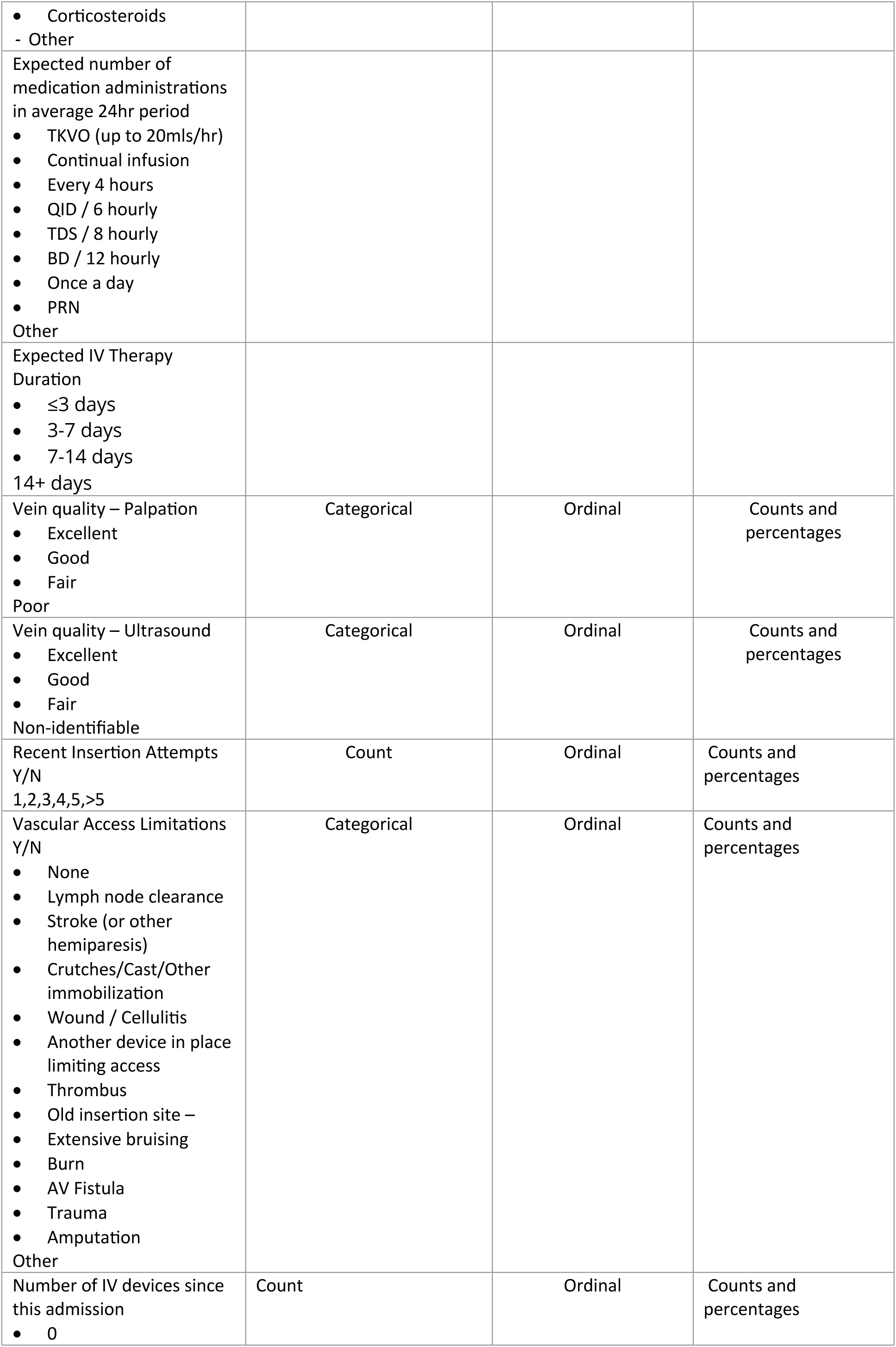
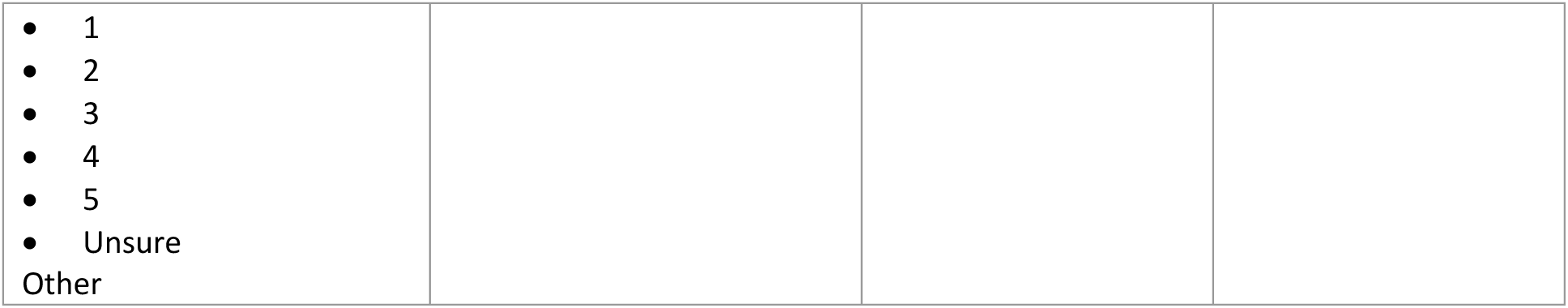

### 4.3 Planned analysis of the primary outcome

The number and proportion of participants that experience device failure (binary, yes/no) will be calculated per group. The between-group difference will be analysed using a generalized linear model with a binomial distribution and the identity link function. The result will be presented as a risk difference (95% CI); *P* value. The primary outcome will be analysed with intention-to-treat, with per-protocol as a sensitivity analysis.

#### Supplementary analysis

The number and proportion of devices that experience failure due to all cause failure will be calculated per group and presented as mean (SD) incidence rate per 1000 hours at risk. This outcome will be compared between-groups as a time-to-event outcome using Cox regression. The regression model will include study group as a fixed effect, with the results presented as a hazard ratio (95% CI); *P* value.

### 4.4 Planned analysis of the secondary outcomes

P values will not be reported for secondary outcomes as these will be exploratory analyses. All secondary outcomes that were composite variables for the primary outcome will be analysed with intention-to-treat, with per-protocol as a sensitivity analysis. They will also be analysed as time-to-event for a supplementary analysis.

#### 4.4.1 Infiltration/extravasation

The number and proportion of participants that experience this outcome (binary, yes/no) will be calculated per group. The between-group difference will be analysed using logistic regression, with study group as the main effect. The result will be presented as an odds ratio (95% CI).

#### 4.4.2 Blockage/occlusion

The number and proportion of participants that experience this outcome (binary, yes/no) will be calculated per group. The between-group difference will be analysed using logistic regression, with study group as the main effect. The result will be presented as an odds ratio (95% CI). This secondary outcome will be analysed with intention-to-treat, with per-protocol as a sensitivity analysis.

#### 4.4.3 Phlebitis

The number and proportion of participants that experience this outcome (binary, yes/no) will be calculated per group. The between-group difference will be analysed using logistic regression, with study group as the main effect. The result will be presented as an odds ratio (95% CI). This secondary outcome will be analysed with intention-to-treat, with per-protocol as a sensitivity analysis.

#### 4.4.4 Thrombosis

The number and proportion of participants that experience this outcome (binary, yes/no) will be calculated per group. The between-group difference will be analysed using logistic regression, with study group as the main effect. The result will be presented as an odds ratio (95% CI). This secondary outcome will be analysed with intention-to-treat, with per-protocol as a sensitivity analysis.

#### 4.4.5 Dislodgment

The number and proportion of participants that experience this outcome (binary, yes/no) will be calculated per group. The between-group difference will be analysed using logistic regression, with study group as the main effect. The result will be presented as an odds ratio (95% CI). This secondary outcome will be analysed with intention-to-treat, with per-protocol as a sensitivity analysis.

#### 4.4.6 Infection

The number and proportion of participants that experience this outcome (binary, yes/no) will be calculated per group. The between-group difference will be analysed using logistic regression, with study group as the main effect. The result will be presented as an odds ratio (95% CI). This secondary outcome will be analysed with intention-to-treat, with per-protocol as a sensitivity analysis.

#### 4.4.7 First insertion success

The number and proportion of participants that experience first insertion success (binary, yes/no) will be calculated per group. The between-group difference will be analysed using logistic regression, with study group as the main effect. The result will be presented as an odds ratio (95% CI).

#### 4.4.8 Number of failed insertion attempts

The counts and totals of failed insertion attempts for each participant *after* randomisation will be calculated per group. The between-group differences will be analysed using an ordinal logistic regression, with study group as the main effect. The result will be presented as an odds ratio (95%CI).

The same analysis and results as above will be presented for number of failed insertion attempts *before* randomisation.

#### 4.4.9 Peripherally placed IV device dwell-time (hours) from device insertion to removal

The mean and SD of the dwell time (hours) will be calculated per group. The between-group difference in dwell time will be computed using linear regression, with study group as the main effect. The result will be presented as mean difference (95% CI).

If the normality assumption of linear regression is not met, the median and IQR of the dwell time will be calculated per group, and a quantile regression will be performed with study group as the main effect. The result will be presented as median difference (95% CI).

#### 4.4.10 Failed Insertion

The number and proportion of participants that received any device after randomisation (binary, yes/no) will be calculated per group. The between-group difference will be analysed using logistic regression, with study group as the main effect. The result will be presented as an odds ratio (95% CI).

#### 4.4.11 Number of devices needed to complete IV treatment

The counts and totals of devices needed for each participant that are inserted after randomisation will be calculated per group. The between-group differences will be analysed using an ordinal logistic regression, with study group as the main effect. The result will be presented as an odds ratio (95%CI).

#### 4.4.12 Type of devices needed to complete IV treatment

The counts of devices for each type for all participants will be calculated per group. The between-group differences will be analysed using a multinomial logistic regression, with study group as the main effect. The result will be presented as a relative risk ratio (95%CI).

#### 4.4.13 Patient-reported pain at device insertion

Rating scales will be treated as a continuous variable and the mean and SD of the ratings will be calculated per group. The between-group difference will be computed using linear regression, with study group as the main effect. The result will be presented as mean difference (95% CI). If assumptions for the linear model are not met, we will use non-parametric median regression instead.

#### 4.4.14 Patient-reported satisfaction with IV device

Rating scales will be treated as a continuous variable and the mean and SD of the ratings will be calculated per group. The between-group difference will be computed using linear regression, with study group as the main effect. The result will be presented as mean difference (95% CI). If assumptions for the linear model are not met, we will use non-parametric median regression instead.

### 4.5 Sensitivity analysis

A sensitivity analysis will be conducted for the primary outcome – DIVA status and difficulty in IV access. The number and proportion of participants that experience device failure (binary, yes/no) will be calculated per group for each analysis. The between-group difference will be analysed using logistic regression, with study group as the main effect. The result will be presented as an odds ratio (95% CI); P value.

### 5.0 Trial status

Recruitment began: 17/07/2023

Recruitment ended:14/02/2024 (last randomisation) Final date of data collection 20/02/2024

SAP signed off: 20/01/2025

Data analysis began: 01/02/2025

## Data Availability

All data produced in the present study are available upon reasonable request to the authors

## List of abbreviations

IQR: interquartile range
IV: intravenous
MC: midline catheter
PICC: peripherally inserted central catheter
PIVC: peripheral intravenous catheter
RCT: randomised controlled trial
ReN: research nurse
SAP: statistical analysis plan
SD: standard deviation
VAS: vascular access specialist

## 7.0 Appendix

**Table 1.**
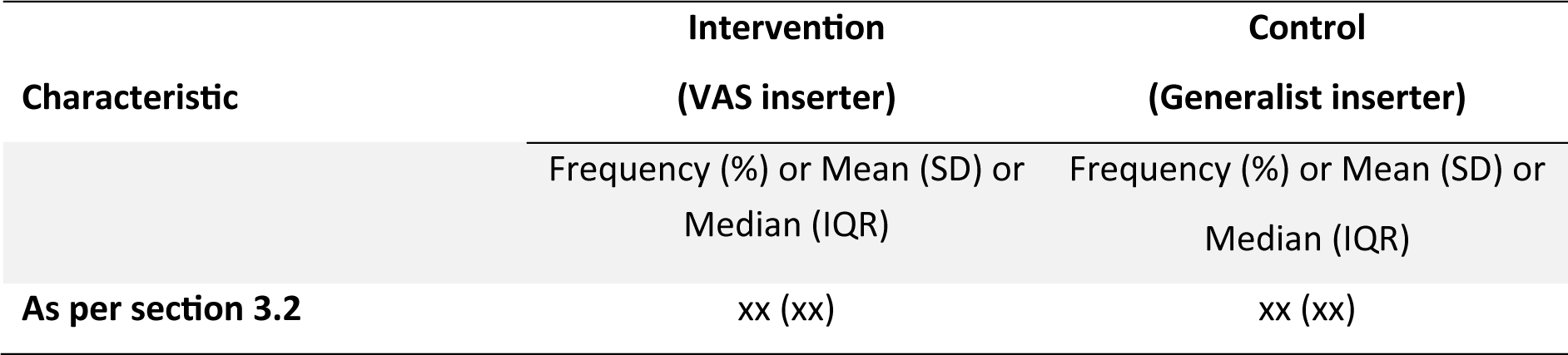
Participant characteristics at baseline.

**Table 2.**
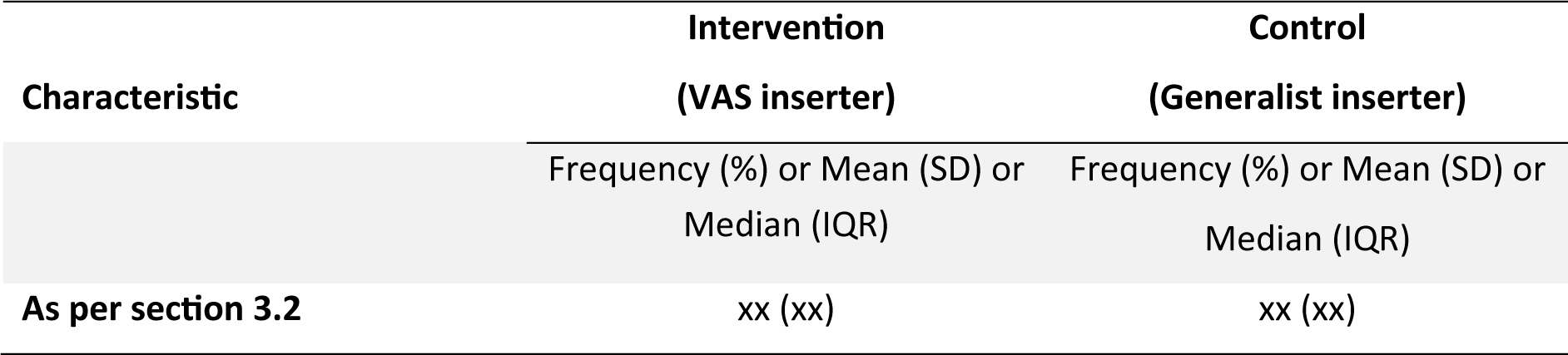
Device characteristics.

**Table 3.**
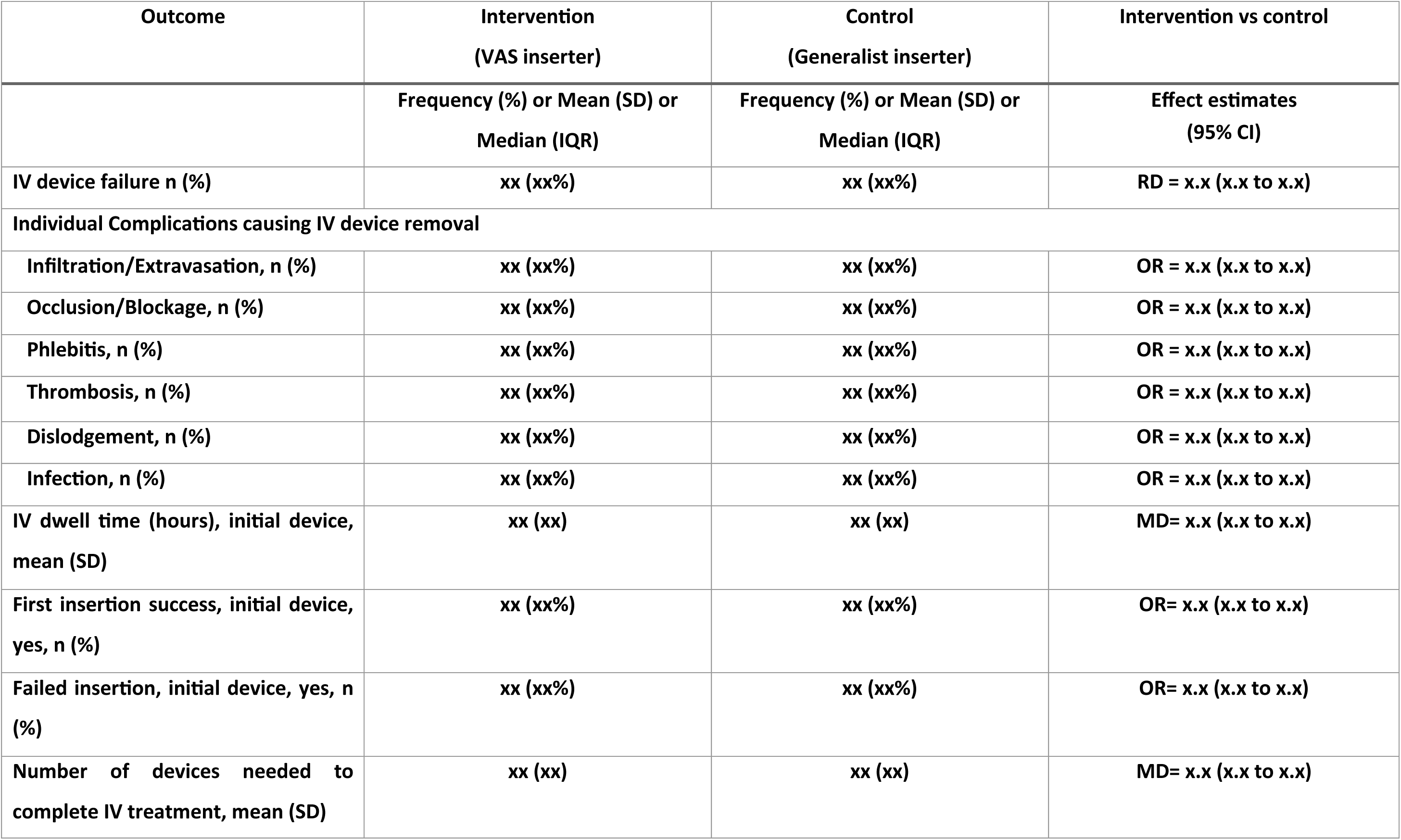
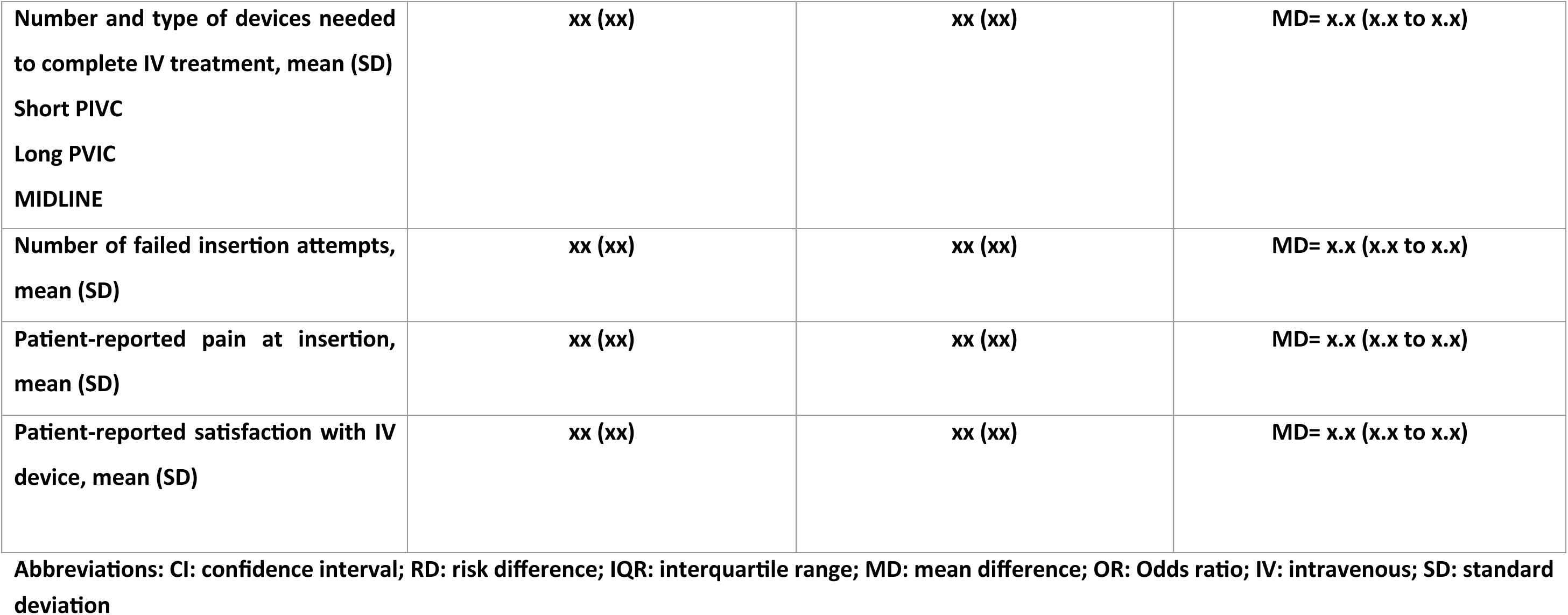
Association between study interventions and study outcomes.

**Supplementary Table 1:**
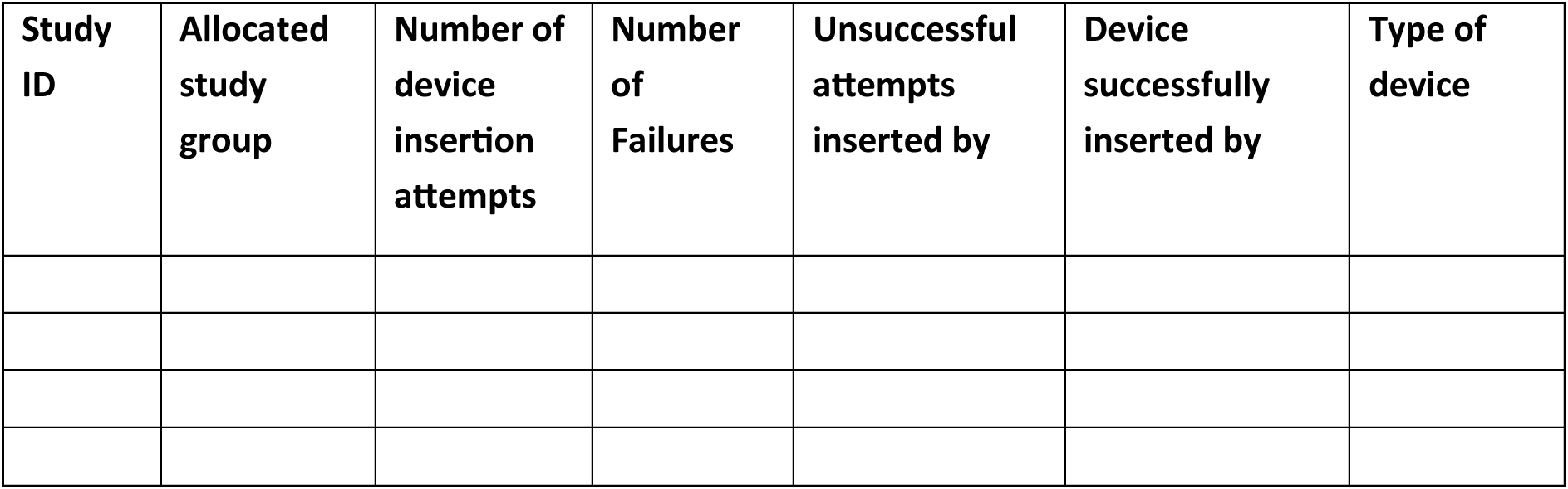
Multiple Device Insertion Attempts.

**Supplementary Table 2:**
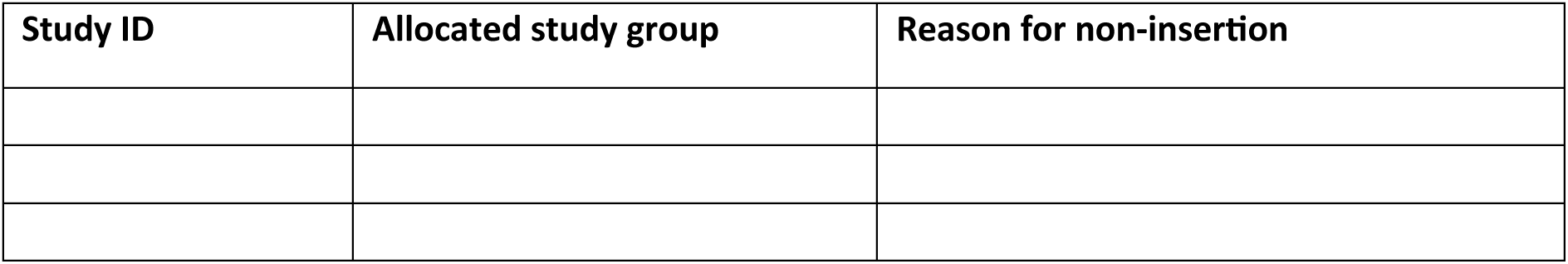
Patients Who Did Not Receive the Device and reason.

**SUPPLEMENTARY Table 3.** Incidence rates for Individual Complications causing IV device removal.

| Outcome | Intervention (VAS)<br>Incidence rate per 1,000 catheter days, mean (95%CI) | Control (Generalist)<br>Incidence rate per 1,000 catheter days, mean (95%CI) | Hazard Ratio (95% CI) |
| --- | --- | --- | --- |
| **Infiltration/Extravasation** | x.x (x.x to x.x) | x.x (x.x to x.x) | HR= x.x (x.x to x.x) |
| **Occlusion/Blockage** | x.x (x.x to x.x) | x.x (x.x to x.x) | HR= x.x (x.x to x.x) |
| **Phlebitis** | x.x (x.x to x.x) | x.x (x.x to x.x) | HR= x.x (x.x to x.x) |
| **Thrombosis** | x.x (x.x to x.x) | x.x (x.x to x.x) | HR= x.x (x.x to x.x) |
| **Dislodgement** | x.x (x.x to x.x) | x.x (x.x to x.x) | HR= x.x (x.x to x.x) |

**Abbreviations: CI: confidence interval; HR: hazard ratio**

## Notes

### Competing Interest Statement

The authors have declared no competing interest.

### Clinical Trial

The trial has been registered with the Australian New Zealand Clinical Trials Registry (ACTRN12623000489695)

### Clinical Protocols

https://www.anzctr.org.au/Trial/Registration/TrialReview.aspx?id=385739&isReview=true

### Funding Statement

This study was funded by the Royal Brisbane and Women Hospital and Royal Brisbane Hospital Foundation(RBWH-PG-00015).The funders have had no input into study design or conduct, data acquisition or analysis, or manuscript preparation

### Author Declarations

Metro North Health Human Research Ethics Committee approval: HREC/2023/MNHA/91818 Griffith University Human Research Ethics Committee approval: 2023/439

